# Connecting BCG Vaccination and COVID-19: Additional Data

**DOI:** 10.1101/2020.04.07.20053272

**Authors:** Devi Dayal, Saniya Gupta

**Affiliations:** Department of Pediatrics, Postgraduate Institute of Medical Education and Research, Chandigarh

**Author notes:** **Correspondence** Dr Devi Dayal, Endocrinology and Diabetes Unit, Department of Pediatrics, 3108, Level III, Advanced Pediatrics Center, Postgraduate Institute of Medical Education and Research, Chandigarh- 160012, India., (O) (R) (M); 2745078. **Sources of funding:** None. **Conflict of interest:** None.

**Keywords:** COVID-19, BCG vaccination, case fatality ratio, mortality, low resource countries

## Abstract

The reasons for a wide variation in severity of coronavirus disease 2019 (COVID-19) across the affected countries of the world are not known. Two recent studies have suggested a link between the BCG vaccination policy and the morbidity and mortality due to COVID-19. In the present study we compared the impact of COVID-19 in terms of case fatality rates (CFR) between countries with high disease burden and those with BCG revaccination policies presuming that revaccination practices would have provided added protection to the population against severe COVID-19. We found a significant difference in the CFR between the two groups of countries. Our data further supports the view that universal BCG vaccination has a protective effect on the course of COVID-19 probably preventing progression to severe disease and death. Clinical trials of BCG vaccine are urgently needed to establish its beneficial role in COVID-19 as suggested by the epidemiological data, especially in countries without a universal BCG vaccination policy.

## Introduction

The ongoing pandemic of coronavirus disease 2019 (COVID-19) caused by severe acute respiratory syndrome coronavirus 2 (SARS-CoV-2) is posing a grave threat to global public health. With more than one million confirmed cases and the global death toll alarmingly crossing 50,000 on April 3, there is a climate of fear, social disruption, institutional breakdown, and scientific uncertainty. The devastation caused by COVID-19 pandemic probably parallels the “Spanish flu” pandemic, still considered the most devastating in human history that affected an estimated one third of the world population at that time of which more than 2.5% died (1). The world governments working in tandem with the health organisations are making all possible efforts to establish countermeasures to reduce the devastating effects of COVID-19. The world’s top healthcare systems appear shaken and the pandemic has forced the medical fraternity to realise something very basic that life is not always in the doctors’ hands, and to feel that being a doctor will never be the same after this pandemic (2). Since there is no definitive treatment or an effective vaccine for disease control, the scientific community has been making frenetic efforts to find measures for cure or for at least reducing the morbidity due to COVID-19 (3). A large number of therapeutic measures such as low-dose methylprednisolone, chloroquine phosphate, hydroxychloroquine, ribavirin, angiotensin receptor 2 blockers, anti-ageing drugs such as azithromycin, quercetin, rapamycin and doxycycline, IL-1 or IL-1R suppressors, protease inhibitors such as lopinavir/ritonavir, acetazolamide, nifedipine, phosphodiesterase inhibitors, convalescent plasma transfusions, tocilizumab, traditional Chinese medicine, statins, zinc supplements, and therapeutic neutralizing and monoclonal antibodies have been either tried or suggested for patients with COVID-19 over a short span of about 4 months in the desperate hope of an effective treatment of these patients (3). Commensurate with the unprecedented times, the World Health Organisation (WHO) has rightly responded by accelerating research in diagnostics, vaccines and therapeutics and called for studying even the unregistered and experimental interventions as formal clinical trials to rapidly establish their safety, efficacy, risks, and benefits (4). In another unprecedented development, hundreds of scientists, physicians, funders, and policy makers from all the major continents of the world have come together to form the COVID-19 Clinical Research Coalition to support WHO’s efforts to counter the COVID-19 pandemic (5). It is in this context that the two recently published epidemiological studies by Miller A et al and Hegarty PK et al, on a correlation between universal Bacille Calmette-Guérin (BCG) vaccination policy and reduced morbidity and mortality for COVID-19, offer a ray of hope for low resource countries (6, 7).

## Methods

To further augment the observations of the two published studies, we hypothesized that countries which adopted a BCG revaccination policy in the past might be at an advantage as the revaccination practices would have sustained the protection offered by BCG vaccination. We specifically compared the case fatality rates (CFR) between the countries with high COVID-19 burden and the countries which had followed BCG revaccination policies in the past. According to the BCG atlas published in 2011, 16 countries adopted BCG revaccination policy in the past (8). Out of these, 4 were excluded from the analysis due either to non-availability of COVID-19 information or the non-specification of the age at second BCG dose. The data on population was taken from the World Population Clock (9). The information on the number of COVID-19 cases and deaths was retrieved from the WHO’s situation report for the day (10).

## Results

We observed a marked difference in the mean CFR between the two groups of countries (5.2% versus 0.6%, p value <0.0001) (Table 1).

**Table 1:** Proportion of COVID-19 affected population and deaths in countries with high disease burden and those with BCG revaccination policy in the past, as on March 29, 2020.

| High burden countries |  |  |  |  |  | Countries with past BCG revaccination practices |  |  |  |  |  |
| --- | --- | --- | --- | --- | --- | --- | --- | --- | --- | --- | --- |
| Country | Population | Total cases | % | Deaths | % | Country# | Population | Total cases | % | Deaths | % |
| China | 1,439,323,776 | 82356 | 0.006 | 3306 | 4.01 | Armenia | 2,963,243 | 424 | 0.014 | 3 | 0.70 |
| Italy | 60,461,826 | 92472 | 0.153 | 10023 | 10.8 | Belarus* | 9,449,323 | 94 | 0.001 | 0 | 0.00 |
| USA | 331,002,651 | 103321 | 0.031 | 1668 | 1.61 | Croatia | 4,105,267 | 657 | 0.016 | 5 | 0.76 |
| Iran | 83,992,949 | 35408 | 0.042 | 2517 | 7.10 | Czech R | 10,708,981 | 2663 | 0.025 | 11 | 0.41 |
| Spain | 46,754,778 | 72248 | 0.155 | 5690 | 7.87 | Fiji | 896,445 | 5 | 0.001 | 0 | 0.00 |
| Germany | 83,783,942 | 52547 | 0.063 | 389 | 0.74 | Kazakhstan* | 18,776,707 | 265 | 0.001 | 1 | 0.37 |
| France | 65,273,511 | 37145 | 0.057 | 2311 | 6.22 | Macedonia | 2,083,374 | 241 | 0.012 | 4 | 1.66 |
| RO Korea | 51,269,185 | 9583 | 0.019 | 152 | 1.58 | Moldova | 4,033,963 | 231 | 0.006 | 2 | 0.86 |
| UK | 67,886,011 | 17093 | 0.025 | 1019 | 5.96 | Norway | 5,421,241 | 3845 | 0.071 | 20 | 0.52 |
| Switzerland | 8,654,622 | 13152 | 0.152 | 235 | 1.78 | Ukraine | 43,733,762 | 418 | 0.001 | 9 | 2.15 |
| Netherlands | 17,134,872 | 9762 | 0.057 | 639 | 6.54 | Uzbekistan* | 33,469,203 | 133 | 0.004 | 2 | 1.50 |
| Belgium | 11,589,623 | 9134 | 0.079 | 353 | 3.86 | Russia | 145,934,462 | 1534 | 0.001 | 8 | 0.52 |
| **Total** | 2,267,127,746 | 534221 | 0.024 | 28302 | 5.29 | **Total** | 281,575,971 | 10510 | 0.004 | 65 | 0.61 |

## Discussion

Our data further supports the observations of previous two studies by Miller A et al and Hegarty PK, of reduced mortality due to COVID-19 in countries with universal BCG vaccination policy. Miller A et al, while rapidly synthesizing the evidence, compared the BCG vaccination policies of a large number of countries with the morbidity and mortality for COVID-19 (6). They rightly chose mortality rates as outcome as the number of reported cases may be influenced by the testing capabilities and underreporting in the low income countries. Confounders such as the country’s standard of medical care and underreporting were adequately excluded. The conclusions drawn from the study that universal BCG vaccination policy is correlated with reduced mortality rates due to COVID-19, thus appear trustworthy. Another conclusion that the countries that established a universal BCG policy earlier had a reduced mortality rate is also reassuring especially for low and middle income economies. The other study by Hegarty PK et al, also arrived at a similar conclusion that countries with national program of universal BCG vaccination appear to have a lower incidence and death rate from COVID-19 (7). Both studies recommend further testing of the hypothesis that BCG vaccine probably offers protection against COVID-19, through randomized controlled trials to determine how fast a BCG induced protective immune response to COVID-19 develops.

The protection offered by BCG vaccine to SARS-CoV-2 has been attributed to its non-specific effects (NSEs) (11). The range of NSEs include a reduction in the incidence of respiratory tract infections in children, antiviral effects and reduced viremia in experimental animals (11). For several viruses such as respiratory syncytial virus, yellow fever, herpes simplex virus and human papilloma virus, a favourable in vitro or in vivo effect has indeed been observed in various studies (11). Based on the hypothesis that BCG vaccination may induce (partial) protection against susceptibility to and/or severity of SARS-CoV-2 infection even during the epidemic, some countries like Australia and the Netherlands which do not practice routine BCG vaccination have been really quick to launch phase 3 clinical trials (12, 13).

A limitation of our study was the calculation of CFR by dividing the number of known deaths by the number of confirmed cases which may not represent the true CFR during an ongoing epidemic (14). We chose this calculation as both the numerator and denominator were rapidly changing and the use was meant only for comparison and not for projecting the mortality due to COVID-19. We also wish to caution against oversimplification of interpretation of BCG vaccination link with COVID-19 as this is derived from the epidemiological data that needs to be confirmed by well designed clinical trials. Furthermore, the CFR depends on multiple factors, in particular, on the ability of the national governments and the healthcare systems to respond to the epidemic. As COVID-19 gains further foothold in low resource countries, the CFR may change significantly and the current conclusions may then become invalid.

Several low resource countries such as India practice universal BCG vaccination policy established since mid-twentieth century. Unlike developed countries where the tuberculosis rates in the population declined significantly prompting their policy makers to adopt a target BCG vaccination approach for high risk populations, the universal vaccination strategy has continued in several high-TB-burden countries (8). The observations of the two recent epidemiological studies alongwith our own data offer hope for a reduced impact of COVID-19 for countries which practice universal BCG vaccination policy, similar to the recently published experience on COVID-19 in children that has instilled some optimism amongst pediatric fraternity (15, 16). Several of these countries have scarce financial and manpower resources to fight the pandemic.

## Data Availability

Yes all data will be made available

